# Endometrial Receptivity on Assisted Reproductive Technology Cycles – a correlation between Serial Serum Estradiol levels and Serial Endometrial Volume Assessment

**DOI:** 10.1101/2020.11.25.20238816

**Authors:** R. Silva Martins, A. Helio Oliani, D. Vaz Oliani, J. Martinez de Oliveira

## Abstract

**Research Question:** Diagnosis of endometrial receptivity is still unclear and conflicting. Despite advances in embryo development during assisted reproductive technologies (ART) cycles, the intricate process of implantation is still matter for debate and research.

**Design:** Serial serum estradiol levels and 3D transvaginal ultrasound performed in women on ART cycle to evaluate a pattern that better predicts implantation rates. 169 subjects on a prospective case control study were assessed. Serial biochemical and biophysical parameters were assessed during ovarian controlled stimulation and results compared in terms of pregnancy outcome.

**Results:** No statistical difference was noted between the two groups in terms of demographics and ART procedures and scores. Serum estradiol was significantly higher in the positive group from day 8 after ovarian controlled stimulation. Endometrial volume and adjusted endometrial volume were significantly higher in the positive group as soon as day 6 of ovarian controlled stimulation.

**Conclusions:** Continuous serum estradiol and 3D endometrial volume and adjusted endometrial volumes may reflect endometrial changes during ART procedures and provide a useful real time tool for clinicians in predicting endometrial receptivity.

## Introduction

The impact of serum estradiol levels in assisted reproductive techniques (ART), has been debated for over 25 years with conflicting results about the effect of supraphysiological levels of estradiol during controlled ovarian stimulation. Some studies showed a negative impact, while others showed a positive impact in ART outcome. The majority showed no impact. (1-5)

A metanalysis was conducted in 2019 and no quality evidence was found to support or refute the value of estradiol levels on the day of hCG administration, as a predictor of pregnancy in ART cycles. (6) Conflicting results may relate to the difference between the way the trials were conducted, the difference of stimulation protocol, number of embryos transferred, and the definition of outcome in terms of pregnancy rates.

According to Paulson (2011) the supraphysiological elevation of serum estradiol compromises endometrial receptivity. (7) This elevation plays a definite role in embryo implantation which is claimed to be dose dependent. (8) As a result of multiple follicle maturation, the rise of serum estradiol to supraphysiological levels, alters endometrial receptivity by morphological and biochemical changes produced against this tissue. (Simon et al.) (9-10)

Still clinical trials, contrast with these claims with conflicting results. By Sharara & McClamrock 1999, (11) no significant impact on implantation rates were reported due to the supraphysiological levels of estradiol; whereas Mirkin et al., 2005 reports that elevated serum estradiol levels have a negative impact on endometrial receptivity especially in fresh embryo transfer cycles. (12)

In a similar way, ultrasonographic parameters have been attempted to understand endometrial receptivity. Still the results of all these features remain uncertain and also conflicting. Ultrasound can assess endometrium changes during a stimulated ART cycle, in a non-invasive manner. Monitoring both follicle development and endometrium during an ART cycle is normal clinical procedure. Many published studies have conflicting results on this subject but the common feature in all, is the lack of continuity on the endometrial assessment. (13-16)

The primary objective of this study is to assess both parameters not only in a single scope pre-determined moment but with a serial prospective continuous evaluation. Endometrium is a responsive tissue that has to undergo serial transformations as a result of the ovarian stimulation. The main goal is to determine the changes and follow up the way both serum estradiol and ultrasound parameters influence this process.

## Material and Methods

Prospective case control study of 169 women in ART cycles of diagnosed infertile couples on ART treatment at our institution during a 2-year period (from January 2017 to December 2018).

Canceled treatments prior to oocyte pickup; cycles with donated gametes; cryopreserved oocyte treatments; cycles for genetic disease screening and embryo selection; cycles with missing or erroneous data; and cycles with elective single embryo transfer were excluded.

The primary data source for this study was the local databases routinely used in the participating centre in ongoing treatments. The data output was anonymized in the extraction for statistical treatment purposes. All data collected and written informed consent was obtained according to the Ethics Committee of our Institution.

Only subjects with viable good grade embryos for transfer (double cleaved embryos) were selected. All subjects have been in a short protocol regimen with antagonist for ovarian controlled stimulation using gonadotropins. All used recombinant human chorionic gonadotropin hormone (rhCG) for induction of ovulation 36 hours prior to oocyte pick up.

Demographics data was collected for all patients and serial serum estradiol levels obtained and ultrasound analysis were performed using the same protocol for all participating subjects. Biochemical and biophysical markers were obtained in all evaluations on a serial continuous evaluation from basal moment (prior to ovarian controlled stimulation) and throughout ovarian controlled stimulation.

Serum levels of estradiol were measured by chemiluminescent enzyme immunoassays (IMMULITE; Diagnostic Products, Los Angeles, CA, USA). Inter-assay and intra-assay coefficients of variation were 9.8% and 9.4%.

Endometrial volume calculation by 3D ultrasound presented as voxels and geometric information of surfaces in a 3D dataset. The results obtained are then converted to millilitres. Adjusted Endometrial volume was also obtained as a ratio between endometrial volume calculated on 3D analysis and uterine volume based on 3D volumetric acquisitions which then generated an estimated uterine volume (also in millilitres).

At day 12 after successful embryo transfer, human gonadochorionic sub-unit B serum levels were obtained. All positive results were confirmed 2 weeks later with the presence of at least one embryo with fetal heart activity.

All data collected was analysed between these two set groups and compared.

Data was analysed in Excel 2019 (Microsoft Corp, Redmond, WA) and IBM SPSS statistics v25 (IBM Corp. Armonk, NY). Continuous variables were analysed with Levene’s test (equality of variances) and visual assessment of the histogram (normality).

Results were expressed as mean±SD, frequency, and percentages. Categorical characteristics of patients were compared with χ2 test. Independent Samples T test and Mann Whitney U tests were used for comparison of numeric variables. For analysis of parametric continuous variables, a t-student test for independent samples was used. Chi-square and Fisher’s exact tests were used to analyse associations between categorical variables. Endometrial thickness, endometrial volume and adjusted endometrial volume were analysed using analysis of variance for repeated measurement data.

Value of p <.05 was considered statistically significant.

Receiver operating characteristic (ROC) curve analysis and comparison of area under curves (AUC) were performed to determine cut-off values of Estradiol, Endometrial Volume and Adjusted Endometrial Volume for the prediction of positive outcome and to calculate their sensitivity, specificity, positive predictive value (PPV), and negative predictive value (NPV). The standard AUC definitions were as follows: AUC = 1 indicates a perfect test, AUC > 0.9 indicates high accuracy and AUC between 0.7 and 0.9 indicates moderate accuracy.

The authors do not report any conflict of interest.

The study protocol has been approved by the Ethics Committee of our Institution (CHCB 22/2017), in accordance with the relevant guidelines and regulations. This study has been conducted in accordance with legal and regulatory requirements, as well as follow generally accepted research practices described in International Conference Harmonisation (ICH) guidelines, Good Clinical Practices (GCP) and the Declaration of Helsinki.

## Results

Subjects were divided into two groups depending on the value of hCG at Day 12 after embryo transfer and ultrasonographic confirmation of clinical pregnancy: 123 on the negative group (72.8%) and 46 on the positive group (27.2%). Demographics characteristics and ART parameters of the 169 subjects are shown in Table 1 and no statistical difference between the two set groups in terms of demographics and ART parameters was met, especially in terms of overall median number of harvested oocytes per cycle defined as the total number of oocytes harvested during oocyte pick up procedure, and rate of collected metaphase II (MII) oocytes. Also, the mean number of cleaved embryos at day 3 of embryo development, and mean number of blastocysts for cryopreservation showed no significant statistical difference between the two set groups.

**Table 1.**
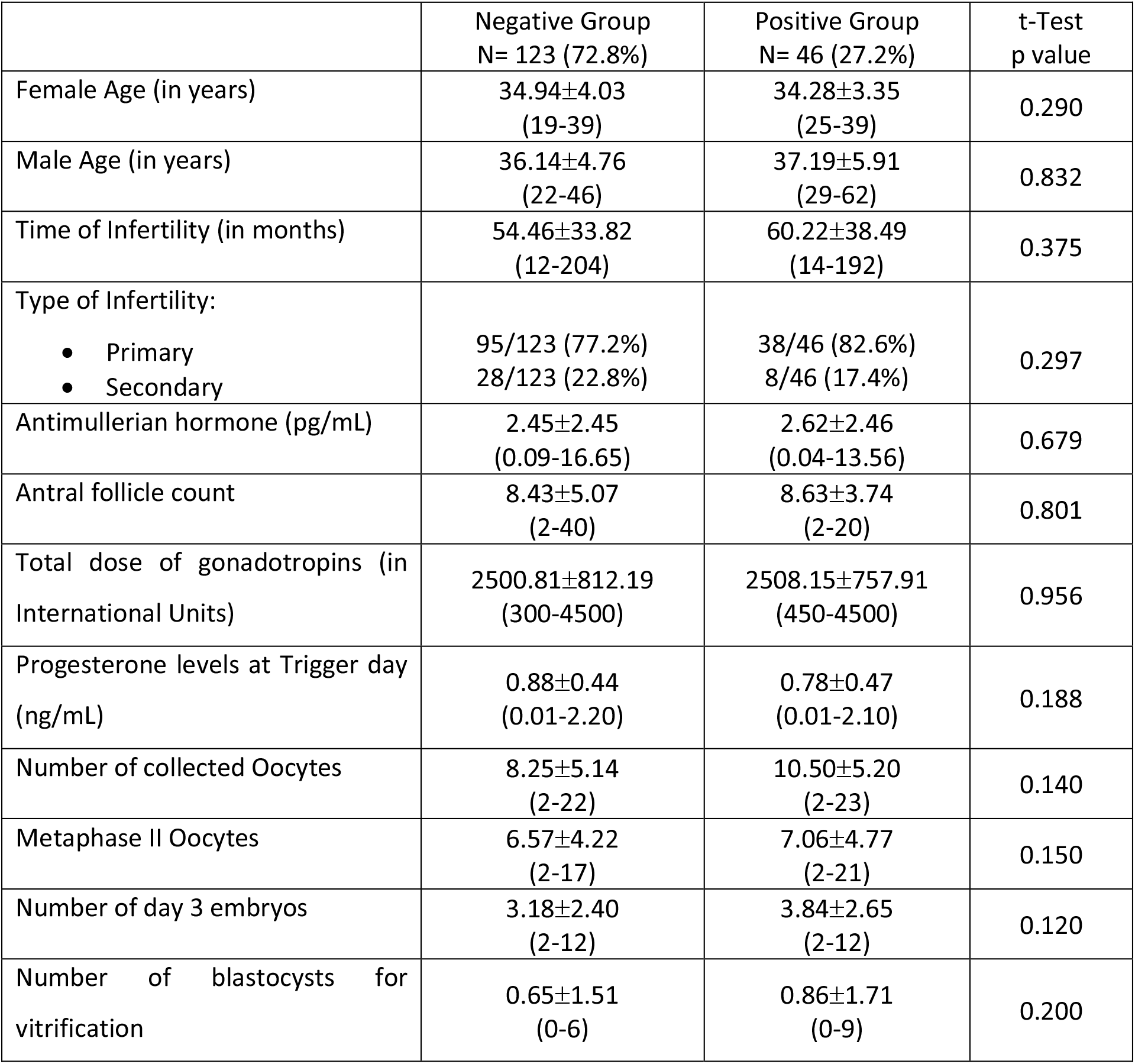
Demographics and ART parameters between two Groups (Positive Group, N = 46 and Negative Group, N = 123). Descriptive statistics between two Groups. Mean values with standard deviation (SD).

|  | Negative Group<br>N= 123 (72.8%) | Positive Group<br>N= 46 (27.2%) | t-Test<br>p value |
| --- | --- | --- | --- |
| Female Age (in years) | 34.94±4.03<br>(19-39) | 34.28±3.35<br>(25-39) | 0.290 |
| Male Age (in years) | 36.14±4.76<br>(22-46) | 37.19±5.91<br>(29-62) | 0.832 |
| Time of Infertility (in months) | 54.46±33.82<br>(12-204) | 60.22±38.49<br>(14-192) | 0.375 |
| Type of Infertility:<br>• Primary<br>• Secondary | 95/123 (77.2%)<br>28/123 (22.8%) | 38/46 (82.6%)<br>8/46 (17.4%) | 0.297 |
| Antimullerian hormone (pg/mL) | 2.45±2.45<br>(0.09-16.65) | 2.62±2.46<br>(0.04-13.56) | 0.679 |
| Antral follicle count | 8.43±5.07<br>(2-40) | 8.63±3.74<br>(2-20) | 0.801 |
| Total dose of gonadotropins (in International Units) | 2500.81±812.19<br>(300-4500) | 2508.15±757.91<br>(450-4500) | 0.956 |
| Progesterone levels at Trigger day (ng/mL) | 0.88±0.44<br>(0.01-2.20) | 0.78±0.47<br>(0.01-2.10) | 0.188 |
| Number of collected Oocytes | 8.25±5.14<br>(2-22) | 10.50±5.20<br>(2-23) | 0.140 |
| Metaphase II Oocytes | 6.57±4.22<br>(2-17) | 7.06±4.77<br>(2-21) | 0.150 |
| Number of day 3 embryos | 3.18±2.40<br>(2-12) | 3.84±2.65<br>(2-12) | 0.120 |
| Number of blastocysts for vitrification | 0.65±1.51<br>(0-6) | 0.86±1.71<br>(0-9) | 0.200 |

Serum estradiol levels on basal (prior to ovarian controlled stimulation) was similar between the two groups. Significantly higher levels were noted on the positive outcome group, but it only met statistical significance at Day 8 after ovarian controlled stimulation. (Table 2) Endometrial Volume and Adjusted Endometrial Volume showed statistical difference from Day 6 after Ovarian controlled stimulation (Table 3). Consistently higher values were seen for both of these biophysical markers on the positive group.

**Table 2.** Serum estradiol measurements between two Groups (Positive Group, N = 46 and Negative Group, N = 123). Mean values with standard deviation (SD).

|  | Negative Group<br>N=123 (72.8%) | Positive Group<br>N= 46 (27.2%) | t-Test<br>p Value |
| --- | --- | --- | --- |
| Basal | 10.47±2.89 | 10.39±2.87 | 0.727 |
| Day 6 after Controlled<br>Ovarian Stimulation | 305.10±130.49 | 381.65±122.46 | 0.055 |
| Day 8 after Controlled<br>Ovarian Stimulation | 840.68±230.49 | 896.32±222.46 | **0.034** |
| Day 10 after Controlled<br>Ovarian Stimulation | 1652.56±607.90 | 1848.65±599.42 | **0.01** |
| Trigger Day with rhCG | 1746.76±654.34 | 2214.65±612.34 | **0.01** |

**Table 3.** Ultrasound parameters between two groups - Endometrial volume and adjusted endometrial volume at baseline, at day 6, 8 and 10 after controlled ovarian stimulation, at trigger day and at embryo transfer day. Ratios in percentages (%) and mean values with standard deviation (SD). rhCG – recombinant human chorionic gonadotropin.

|  |  | Negative Group<br>N=123 (72.8%) | Positive Group<br>N= 46 (27.2%) | t-Test<br>p Value |
| --- | --- | --- | --- | --- |
| Basal | Endometrial Volume<br>(in mm3) | 2.52±0.71 | 2.77±0.63 | 0.54 |
|  | Adjusted Endometrial<br>Volume | 4.60±1.42 | 5.51±1.28 | 0.21 |
| Day 6 after<br>Controlled<br>Ovarian<br>Stimulation | Endometrial Volume<br>(in mm3) | 3.08±0.66 | 3.33±0.57 | **0.024** |
|  | Adjusted Endometrial<br>Volume | 5.63±1.50 | 6.67±1.38 | **0.001** |
| Day 8 after<br>Controlled<br>Ovarian<br>Stimulation | Endometrial Volume<br>(in mm3) | 3.90±0.94 | 4.40±0.71 | **0.002** |
|  | Adjusted Endometrial<br>Volume | 7.28±2.67 | 8.98±2.47 | **0.001** |
| Day 10 after<br>Controlled<br>Ovarian<br>Stimulation | Endometrial Volume<br>(in mm3) | 4.12±1.01 | 4.91±0.82 | **0.001** |
|  | Adjusted Endometrial<br>Volume | 7.60±2.54 | 9.99±2.61 | **0.001** |
| Trigger Day<br>with rhCG | Endometrial Volume<br>(in mm3) | 4.52±1.00 | 5.33±0.76 | **0.001** |
|  | Adjusted Endometrial<br>Volume | 8.30±2.52 | 10.76±2.62 | **0.001** |
| Embryo<br>Transfer Day | Endometrial Volume<br>(in mm3) | 4.84±1.01 | 5.59±0.77 | **0.001** |
|  | Adjusted Endometrial<br>Volume | 8.32±2.58 | 10.83±2.73 | **0.001** |

Receiver operating characteristic (ROC) curve analysis and comparison of area under curves (AUC) were performed for Estradiol, Endometrial Volume and Adjusted Endometrial Volume on the prediction of positive outcome. Values of 0,701; 0,723 and 0,756 were obtained respectively for the examined parameters. (Figure 1)

**Figure 1.**
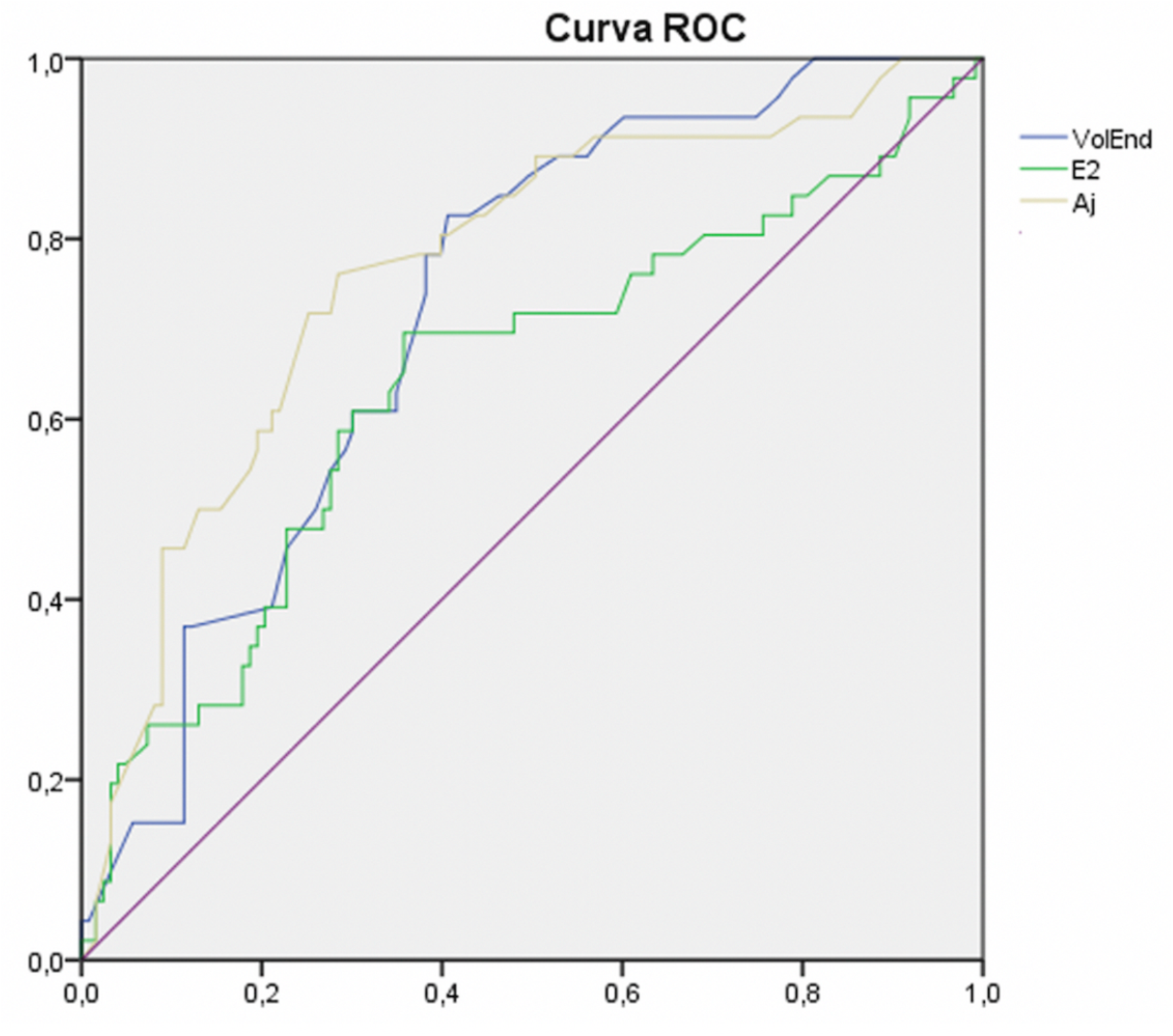
The ROC area under curves for prediction of positive outcome. Vol End – Endometrial Volume; E2 – Serum Estradiol; Aj – Adjusted Endometrial Volume.

In this study the intra-observer reliability was 0.96. In addition, because all measurements were performed by the same operator in this study there was no inter-observer variability.

## Discussion

The process of endometrial transformation from proliferative phase to secretory phase under the steroids hormonal influence, called endometrial decidualization is a set goal for optimal implantation. Cyclic changes of endometrium are regulated by ovarian hormones. (17) This pattern may alter due to the supraphysiological hormonal levels during ART cycles.

Single analysis of endometrial pattern at trigger day has been the most used, with contradictory findings. Recent studies (*Silva Martins, R. et al*.) have proven that perhaps serial evaluations provide better understanding rather than a single scoop at a pre-determined phase of the process. (18-19) The main purpose of this study was to assess both parameters not only in a single scope pre-determined moment but with a serial prospective continuous evaluation.

Conflicting results have been published, but this new methodological approach to endometrial assessment during ART procedures, may shine a new light in better understating the way endometrium transforms and becomes receptive for successful embryo implantation. Serial evaluations, of both biochemical and biophysical parameters, better translate endometrium transformations and may be a base for the understating of endometrial receptivity. (20-21)

In this study we aimed to assess endometrial evolution in order to ascertain a plausible predictive non-invasive diagnostic tool for clinicians to better understand endometrium changes.

Endometrial volume and adjusted endometrial volume proven to be more effective with differences shown since the beginning of ovarian controlled stimulation. Both groups were similar at baseline but as soon as controlled ovarian stimulation started, the differences between those with a positive outcome and those without were clearly met.

We have also been able to show differences between the two groups in terms of endometrial and adjusted endometrial volume in early stages of endometrial development under the influence of controlled ovarian stimulation. Higher volumes were seen in the positive controls, but the changes were more evident in early stages (between day 6 and day 8 of ovarian controlled stimulation for endometrial volume, and between day 8 and day 6 and also between day 10 and day 8 of ovarian controlled stimulation in adjusted endometrial volumes).

In a similar way the rise of serum estradiol was significantly higher in the positive outcome group, despite the fact that the number of oocytes on pick up, mature oocytes and number of cleaved and blastocysts was similar between both groups. This reflects the effects that serum estradiol has directly on endometrium in later phases of its development prior to embryo implantation.

All of these findings may prove to be a useful management tool for clinicians in order to establish yet another diagnostic tool for better decision making in selective embryo transfer. This new methodology uptake, and a new perspective of endometrial analysis is certainly the strongest factor of this study, as well as the number of continuous serial evaluations on the same patient, throughout the ovarian controlled stimulation and its effects on endometrium. We could not refrain to uphold expectation of these results as they show a serial of values, demonstrating a certain pattern of evolution on a transforming living tissue and its natural adaptations to a complex and yet unknown process. Also, the number of subjects in this study constitutes a limitation and further larger studies must be carried out in order to certainly establish this promising results.

## Conclusions

The underlying mechanism that results in failure of implantation of a good quality embryo on a supposed receptive endometrium is still unclear. Endometrial receptivity is still up to this date a controversial subject and a hot topic for discussion and analysis.

Most published studies present conflicting results and are still controversial. In some cases, the diagnosis has been too invasive and lacking reliability especially in women with irregular menstrual cycles. The adaptative changes and continuous evolution of the endometrium makes it difficult to establish a normative pattern of development in way to provide useful information regarding its receptiveness, in real time and in a personalized individualized setting.

Ultrasound developments have been able to clarify and make aware more information about the morphokynetics of this tissue and its changes throughout the cycle. Better understanding of the role that makes an endometrium receptive may be the key in solving these issues, providing a diagnostic tool that will enhance ART cycles and elective embryo transfers more effective in producing better outcomes. Also, the possibility to determine a serial continuous pattern of serum estradiol and its role on endometrial development may provide information useful on determining endometrial receptivity.

This study showed that endometrial volumetry may identify a receptive endometrium as soon as day 6 of ovarian controlled stimulation. Serum Estradiol also showed some predicting value, with statistical significance. Nevertheless, the difference noted between groups did not affect the number of oocytes in oocyte retrieval, nor the number of mature oocytes. Also, the number of cleaved embryos and blastocysts was similar in both groups. This shows that serum estadiol has potential effects on endometrium development in later phase of its development. In this way clinicians may be made aware of this possibility and further enhance its procedures with better knowledge weather or not to perform embryo transfer on that given cycle.

## Data Availability

Encrypted non-disclosure data available at Open Science Framework database for peer review purpose only

https://osf.io/hr25m/?view_only=8d5f6dcb8b25420bbd9188382163e7d7

## List of Abbreviations

ART: Assisted Reproductive Technology
CHCB: Centro Hospitalar Cova da Beira
ER: Endometrial Receptivity
ERA: Endometrial Receptivity Array
GCP: Good Clinical Practices
hCG: human chorionic gonadotropin
ICH: International Conference Harmonisation
IU: International Units
SD: Standard Deviation
WOI: Window of Implantation
2D: Two Dimensions
3D: Three Dimensions

## Declaration Statements

### Ethics approval and consent to participate

The study has been approved by the Ethics Committee of Centro Hospitalar Universitário Cova da Beira (study committee reference number: CHCB 22/2017). Oral and written consent was obtained for all willing participants prior to registering for this study. Patient Informed consent to participate in this study CHCB 22/2017.

### Consent for publication

Not Applicable.

### Availability of data and material

Encrypted non-disclosure data available at Open Science Framework database for peer review purpose only. Project name Physical Biomarkers in Endometrial Receptivity with access link: https://osf.io/hr25m/?view_only=8d5f6dcb8b25420bbd9188382163e7d7

### Author’s Contribution

RSM, AHO and JMO are responsible for conception and study design. RSM has been the principal investigator and the principal responsible for acquisition and analysis of data. RSM has been responsible for data interpretation. DVO, AHO and JMO have been responsible for reviewing the article for publication. All authors have read and approved the manuscript and can be personally accountable for the contributions and can ensure that questions related to the accuracy and integrity of any part of the work.

## Acknowledgements

We would like to appraisal M.S. for the help with the statistical analysis of data.

